# Assessment of the Impact of COVID-19 pandemic on population level interest in Skincare: Evidence from a google trends-based Infodemiology study

**DOI:** 10.1101/2020.11.16.20232868

**Authors:** Hasan Symum, Md. F. Islam, Habsa K. Hiya, Kh M. Ali Sagor

## Abstract

**Background:** COVID-19 pandemic created an unprecedented disruption of daily life including the pattern of skin related treatments in healthcare settings by issuing stay-at-home orders and newly coronaphobia around the world.

**Objective:** This study aimed to evaluate whether there are any significant changes in population interest for skincare during the COVID-19 pandemic.

**Methods:** For the skincare, weekly RSV data were extracted for worldwide and 23 counties between August 1, 2016, and August 31, 2020. Interrupted time-series analysis was conducted as the quasi-experimental approach to evaluate the longitudinal effects of COVID-19 skincare related search queries. For each country, autoregressive integrated moving average (ARIMA) model relative search volume (RSV) time series and then testing multiple periods simultaneously to examine the magnitude of the interruption. Multivariate linear regression was used to estimate the correlation between countries’ relative changes in RSV with COVID-19 confirmed cases/ per 10000 patients and lockdown measures.

**Results:** Out of 23 included countries in our study, 17 showed significantly increased (p<0.01) RSVs during the lockdown period compared with the ARIMA forecasted data. The highest percentage of increments occurs in May and June 2020 in most countries. There was also a significant correlation between lockdown measures and the number of COVID-19 cases with relatives changes in population interests for skincare.

**Conclusion:** Understanding the trend and changes in skincare public interest during COVID-19 may assist health authorities to promote accessible educational information and preventive initiatives regarding skin problems.

## Introduction

Corona Virus Disease 2019 (COVID-19) which was first thought of pneumonia later detected as a novel coronavirus, SARS-CoV-2, on December 31, 2019, originated in Wuhan (Hubei), China. Due to the continuous increment of new cases WHO (World Health Organization) first called it an international emergency on January 31, 2020, then later on March 11, 2020, it was declared as a pandemic. On September 10, 2020, total reported cases were approximately 27.5 million in more than 213 countries and territories around the world with a mortality rate of 4.30% and daily cases of 258,000. To restrict the propagation of new cases, lots of preventive measures (international flight ban, social distancing, mass gathering ban, issuing stay home, etc.) were taken around the world. This restriction brought disruption in the daily life of people as they can’t do physical activity properly. Besides the increase of screen exposure leads the young generation to the sleepless problem.^1^ People of dysmorphic concern usually went to Beaty Service for improving their behavioral appearance through engagement and practice are now having higher dysmorphic concern for “Stay Home” order.^2^ In addition, dermatitis patients who have skin problems like hair and scalp diseases usually needed to take treatment from hospitals now also suffering from this situation.^3^

With the increase of internet users around the globe, Infodemiology has become a very useful informatics method, especially for epidemic and pandemic situations. The Infodemiology study provides real-time surveillance in the health system by Google Trends which has already been used for studying epidemics like the 2009 influenza.^4,5^ Besides, Google Trends also used for monitoring behavioral analysis such as for analyzing the smoking ban in China, incidence analysis of HIV infection from Tweets, and multiple sclerosis (MS) analysis in English-speaking countries.^6–8^ Several studies have done using Google Trends for monitoring the impact of COVID-19 on public interests-based health parameters such as loss-of-smell in the COVID-19 patients, dental problems associated with “toothache” and “tooth pain” keyword search, rapid declination on total joint arthroplasty (TJA), and increase interest in smoking cessation.^9–14^ In addition, Google Trends also used for finding the anxiety associated with search queries terms of “face mask” and “wash hands”.^15^

There have been very few studies that have done about the impact of COVID-19 in the dermatological study using Google Trends. The investigation of people’s interests in dermatologic conditions (i.e. “acne”, “eczema”, “hair loss”, etc.) was the only research where Google Trends were used, specifically, there is no research found about the effect of COVID-19 on skincare.^16^ In our study we aimed to determine whether there is any significant change in the public interest in skincare during COVID-19 by using Google Trends.

## Methods

### COVID-19 Case Data

Daily data on laboratory-confirmed cases and deaths were collected in the COVID-19 web tracker by the Johns Hopkins University Center for Systems Science and Engineering (CSSE).^17^ Daily confirmed COVID-19 cases were retrieved for the worldwide and individual countries from January 22, 2020, to August 31, 2020.

### Country Specific Data

Information on COVID-19 related lockdown restriction and stay home order duration were obtained from each state country’s official website. Weekly seasonally adjusted national and state insured unemployment rates were obtained from the World bank website from January 22, 2020, to August 1, 2020.^18^ Each countries total population for the year 2019 was extracted from the World meter to calculate per 100 thousand COVID-19 cases.

Skin disease-related (per 100 thousand children) in the individual were also included in our study from the national disease burden study for the year 2016. Finally, state-by-state percentage of uninsured children and child immunization rates were obtained from America’s Health Rankings website which provides national health benchmarks and state rankings on various health measures.^19^

### Measures

Country level Inclusion criteria were: a population of more than one million, more than 100 COVID-19 deaths per million, and 50% internet users of the total population. Google Trends country wise data was queried from January 2015 to August 2020 using the search topic “Skin Care”. Daily confirmed COVID-19 cases were obtained from the data repository by the Johns Hopkins University

### Statistical Analysis

Interrupted time-series analysis was conducted as the quasi-experimental approach to evaluate the longitudinal effects of COVID-19 skincare related search queries around the world.^20,21^ For this purpose, this analysis involved identifying the best fit autoregressive integrated moving average (ARIMA) model for each countries’ RSV time series and then testing multiple periods simultaneously to examine the magnitude of the interruption. ARIMA modeling is appropriate for studying the transient and permanent effect of interruption events by taking account of dependency and seasonality between data points in time series.^22,23^ For each country, the four steps Box-Jenkins methodology was used to select the best fitting time series ARIMA model and this approach has been widely used in modeling time series interruptions particularly with health policy changes.^24^ To estimate the magnitude and duration of the effect of COVID-19 related restriction measures, dummy variables were added for every month after March 11, 2020, on which WHO declared COVID-19 as a pandemic. Estimates were made both for the five months combined and for the separate month. Output was measured in the relative changes (%) in RSV to the expected level without the occurrence of COVID-19 restrictions for each period. In addition, we also used a multivariate linear regression model to estimate the correlation between the country’s relative changes in RSV after 11th March with COVID-19 -confirmed cases/ per 10000 patients, lockdown orders, after adjusting covariates of the unemployment growth, skin disease rate and internet user rates. All the statistical analyses were performed using R and a p-value less than 0.01 was considered significant for effect sizes. IRB approval was not required because this study did not involve human subjects.

## Results

Across the world, relative search volume (RSV) for skincare has increased continuously after WHO (World Health Organization) declared COVID-19 as a global pandemic from March 11, 2020, and it continues until May 09, 2020. After that, the rate of RSV was decreased slightly but still, it remains higher in many countries. In our study, we selected 23 countries from all over the world where only two Asian Countries and South Africa met the design criteria along with the countries of Europe and the Americas (Appendix Table-1). Among the selected countries, Chile has the most confirmed COVID-19 cases (20,061) and Belgium has the most confirmed death rate in per hundred thousand population. The lockdown style imposed in most countries was nationwide except five countries where Belgium, Canada, and the USA imposed state-based lockdown, and Chile and Russia imposed city-based lockdown. Of 23 countries, 17 showed significantly (p<0.01) increased RSVs during the lockdown period compared with the ARIMA forecasted data with a mean confidence interval (CI) of 95% (Table 1). The correlation between lockdown measures and the number of COVID-19 cases found significant from the multivariate linear regression analysis with a p-value of less than 0.05(Table 2).

**Table 1:** Monthly relative volume changes for skincare after COVID-19 pandemic.

| Percentage (%) relative search volume changes Mean (95% CI) |  |  |  |  |  |  |
| --- | --- | --- | --- | --- | --- | --- |
| Country | March-2020 | April-2020 | May-2020 | June-2020 | July-2020 | August-2020 |
| Worldwide | -4.13<br>(-8.48,0.22) | 22.48<br>(16.37,28.60) | 29.62<br>(22.23,37.00) | 22.92<br>(14.46,31.38) | 20.62<br>(11.26,29.97) | 16.3<br>(6.08,26.53) |
| Argentina | -2.83<br>(-20.64,14.98) | 23.00<br>(3.34,42.67) | 48.16<br>(26.82,69.49) | 47.99<br>(25.14,70.84) | 73.91<br>(49.67,98.16) | 49.38<br>(23.85,74.92) |
| Belgium | 33.99<br>(16.93,51.05) | 71.47<br>(52.40,90.54) | 66.47<br>(46.51,86.43) | 31.58<br>(11.36,51.79) | 17.91<br>(-3.18,39.00) | 31.75<br>(11.02,52.47) |
| Brazil | 38.63<br>(30.03,47.24) | 87.93<br>(75.36,100.51) | 88.08<br>(73.36,102.80) | 71.53<br>(55.19,87.88) | 75.97<br>(58.14,93.80) | 47.74<br>(28.09,67.40) |
| Canada | -17.03<br>(-22.53,-11.53) | 31.83<br>(26.17,37.50) | 43.77<br>(37.50,50.04) | 40.95<br>(34.50,47.9) | 24.07<br>(18.05,30.10) | 27.24<br>(21.25,33.22) |
| Chile | -2.12<br>(-25.98,21.73) | 77.42<br>(52.07,102.76) | 51.03<br>(28.30,73.76) | 138.37<br>(112.91,163.84) | 131.23<br>(107.22,155.24) | 153.39<br>(129.21,177.58) |
| Colombia | -12.88<br>(-27.73,1.97) | -8.43<br>(-26.98,10.13) | 8.61<br>(-12.85,30.06) | 27.34<br>(3.12,51.57) | 32.52<br>(6.40,58.65) | 37.04<br>(8.72,65.36) |
| Dominican Republic | 53.08<br>(4.01,102.15) | 66.15<br>(15.76,116.53) | 86.68<br>(35.01,138.35) | 66.15<br>(13.22, -119.07) | 81.08<br>(26.93,135.23) | 60.55<br>(5.20,115.89) |
| Ecuador | -9.36<br>(-46.66,27.94) | -1.65<br>(-41.45,38.15) | -1.65<br>(-43.80,40.50) | 63.92<br>(19.54,108.29) | 54.28<br>(7.78,100.77) | 92.84<br>(44.32,141.36) |
| France | -2.40<br>(-17.87,13.07) | 19.00<br>(0.11,37.88) | 25.72<br>(4.31,47.13) | 36.67<br>(11.46,61.87) | 12.89<br>(13.64,39.42) | 7.15<br>(-20.72,35.02) |
| Germany | 35.82<br>(24.50,47.14) | 24.77<br>(13.71,35.83) | 20.56<br>(8.61,32.51) | 19.66<br>(6.50,32.82) | 15.70<br>(2.45,28.96) | 3.22<br>(-10.41,16.86) |
| Italy | 16.64<br>(-1.15,34.43) | 56.55<br>(36.57,76.54) | 49.96<br>(26.19,73.73) | 9.33<br>(-16.80,35.45) | 12.18<br>(-16.42,40.78) | 6.50<br>(-24.18,37.17) |
| Kuwait | 59.29<br>(7.67,110.90) | 35.89<br>(-4.59,76.37) | 36.16<br>(-7.38,79.70) | 39.23<br>(-6.25,84.72) | 8.94<br>(-34.38,52.25) | 10.05<br>(-32.87,52.98) |
| Mexico | -3.34<br>(-17.50,10.83) | 6.42<br>(-8.83,21.67) | 38.36<br>(21.32,55.39) | 62.01<br>(43.51,80.51) | 74.34<br>(54.51,94.17) | 62.13<br>(40.82,83.44) |
| Netherlands | -3.97<br>(-15.56,7.61) | 50.87<br>(38.60,63.15) | 32.46<br>(19.70,45.21) | 19.14<br>(5.06,33.22) | -1.68<br>(-16.19,12.83) | 9.83<br>(-4.81,24.46) |
| Oman | -37.03<br>(-71.30, -2.76) | -15.55<br>(-47.49,16.70) | -18.93<br>(-52.65,14.79) | 55.14<br>(22.33,87.95) | 30.38<br>(-2.12,62.88) | 46.12<br>(11.91,80.34) |
| Panama | -3.89<br>(-57.80,50.02) | 33.07<br>(-27.38,93.52) | 65.10<br>(-1.24,131.45) | 146.42<br>(74.67,218.18) | 47.85<br>(-13.35,31.90) | 84.82<br>(3.31,66.33) |
| Portugal | 3.61<br>(-14.32,21.54) | 58.10<br>(38.82,77.37) | 53.49<br>(32.84,74.14) | 38.18<br>(-16.53,59.83) | 9.28<br>(12.31,57.09) | 60.04<br>(36.28,83.80) |
| Romania | 15.59<br>(-1.30,32.48) | 119.92<br>(102.80,137.05) | 64.82<br>(47.94,81.70) | 47.99<br>(27.62,68.37) | 34.70<br>(-51.06,53.63) | 48.39<br>(26.59,70.18) |
| Russia | -1.99<br>(-22.75,18.77) | -3.06<br>(-28.98,22.86) | -7.32<br>(-37.53,22.89) | -7.32<br>(-41.19,26.64) | -13.71<br>(30.79,71.54) | -19.04<br>(-59.48,21.20) |
| South Africa | 2.38<br>(-11.58,16.34) | 48.09<br>(32.62,63.55) | 82.09<br>(65.21,98.98) | 60.04<br>(41.46,78.63) | 51.16<br>(-18.580,17.17) | 62.06<br>(41.64,82.48) |
| Spain | -21.77<br>(-35.70, -7.84) | 18.69<br>(4.07,33.30) | 23.70<br>(8.43,38.97) | -3.21<br>(-19.42,12.68) | 0.69<br>(-15.80,17.17) | 4.53<br>(-12.53,21.58) |
| UK | 1.77<br>(-6.67,10.22) | 75.56<br>(65.95,85.18) | 80.54<br>(70.39,90.69) | 52.29<br>(41.51,63.06) | 55.50<br>(44.05,66.95) | 30.30<br>(19.47,41.13) |
| USA | -11.70<br>(-17.65, -5.76) | 25.29<br>(17.76,32.82) | 42.63<br>(34.54,50.72) | 30.10<br>(21.35,38.85) | 26.77<br>(18.40,35.13) | 21.61<br>(12.91,30.31) |

**Table 2:**
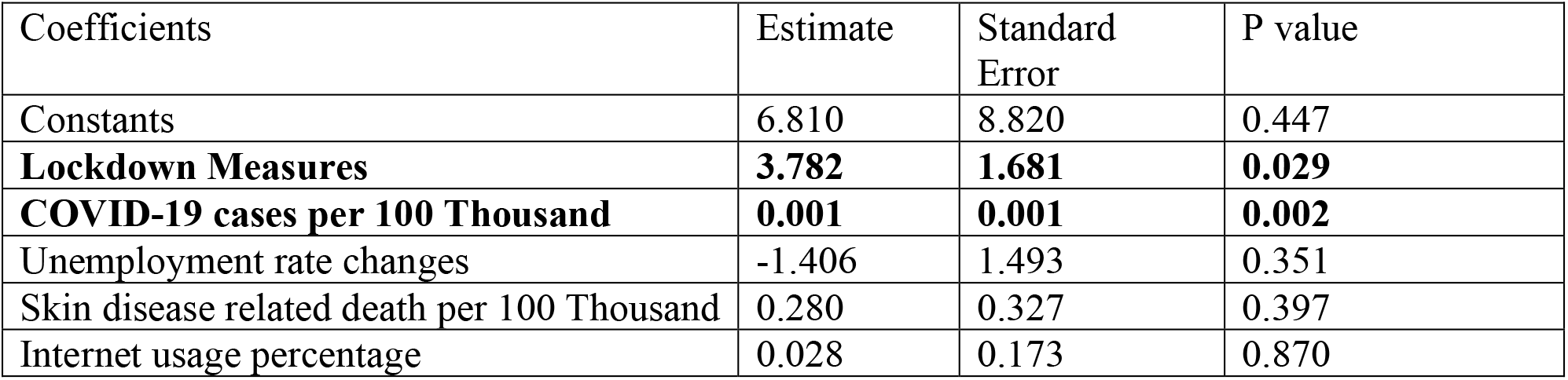
Multi-variate regression results.

The worldwide interest in skincare started to increase from March 15 with a significant increment in April (22.48%, CI of [16.37, 28.60] %) and reached the highest increment in May (29.62%, CI of [22.23, 37.00] %). After May, the increment in worldwide RSVs started to slow down (Figure 1), but the trend is still significantly higher compared to the forecasted value in August (16.3%, CI of [6.08, 26.53] %). The RSV trend in two south Asian countries showed a varied impact during the COVID-19 pandemic and lockdown restriction measures. Compared with the forecasted value, Oman showed a significant increment in RSVs in May and August while there were no such significant changes in Kuwait. Besides, South Africa showed a significant increment in RSVs in March (48.09%, CI of [32.62, 63.55] %) and April (82.09%, CI of [65.21, 98.98] %) though from May the trends started to decrease.

**Figure 1:**
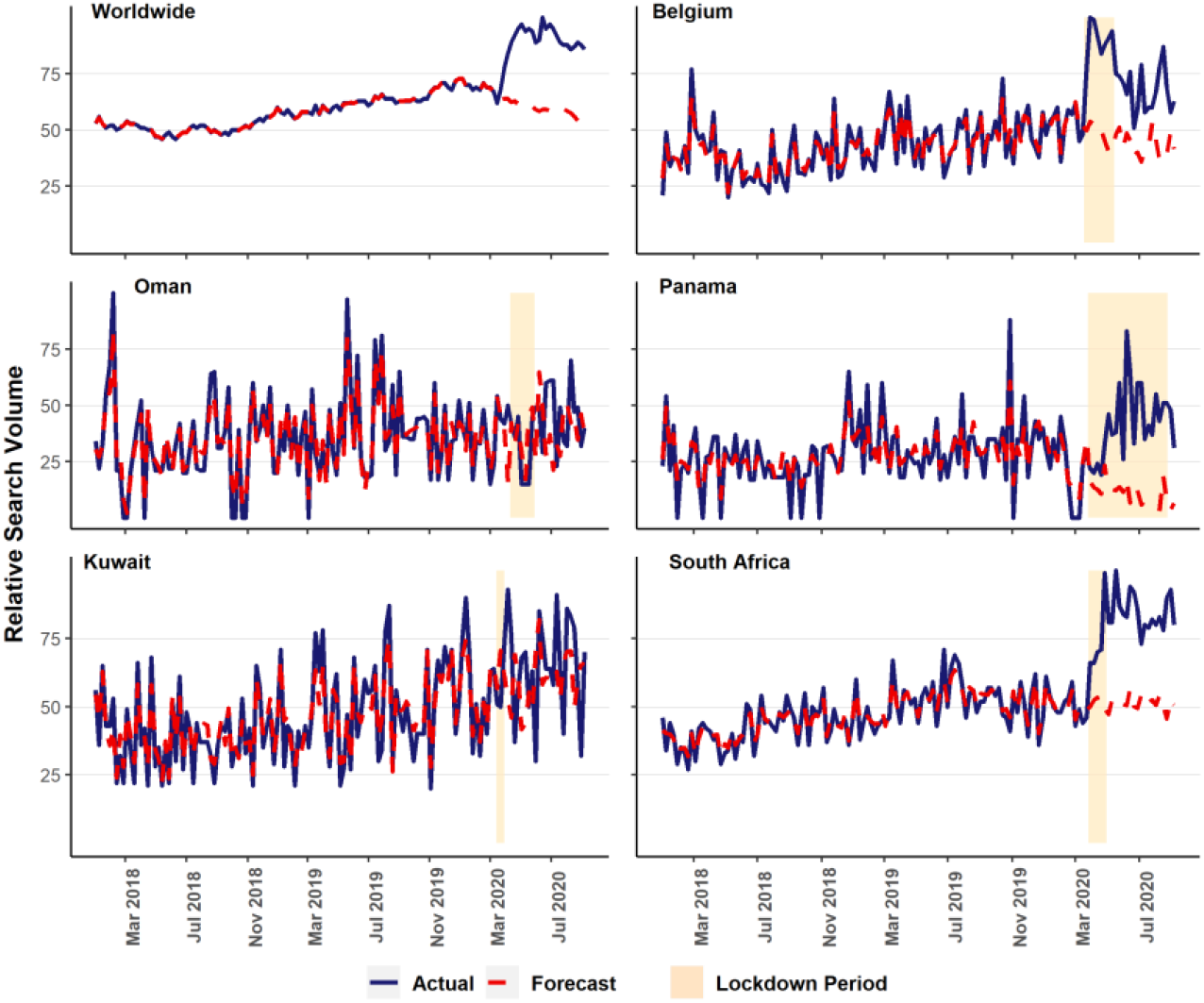
Acutal and ARIMA forecasted mean RSV values in worldwide and five countries.

Among the countries in the Americas, the USA, Brazil, Canada, Chile, and Panama showed the stepper increase in RSVs at the beginning of the lockdown whereas Mexico, Columbia, and Ecuador showed a higher increment in the late period (Figure 2). The highest increment in RSV values for the Americas was found in Chile in August (153.39%, CI of [129.21, 177.58] %). In the USA, the highest percentage change of RSVs seen in May (42.63%), and the trend still above the forecasted trend. The trends of Argentina and the Dominican Republic followed the forecasted trend though there is still an unprecedented stepper increase in RSV values. Of all selected countries in the Americas, only Columbia and Ecuador showed a decrease in RSVs with no significant relationship. Like the Americas, European countries also showed the earlier stepper increase in RSVs with a significant percent increase except for Russia (Figure 3). Italy and Spain showed a significant increase in RSVs at the beginning of lockdown, though in the late part of the lockdown this change started to decrease. The UK had the most unprecedented trend in the RSVs, and the significant percent increase had seen on April 26 (80.54%, CI of [70.39,90.69] %). Although Germany and Spain had shown the earlier increment, the trend went down rapidly after the lockdown withdrew.

**Figure 2:**
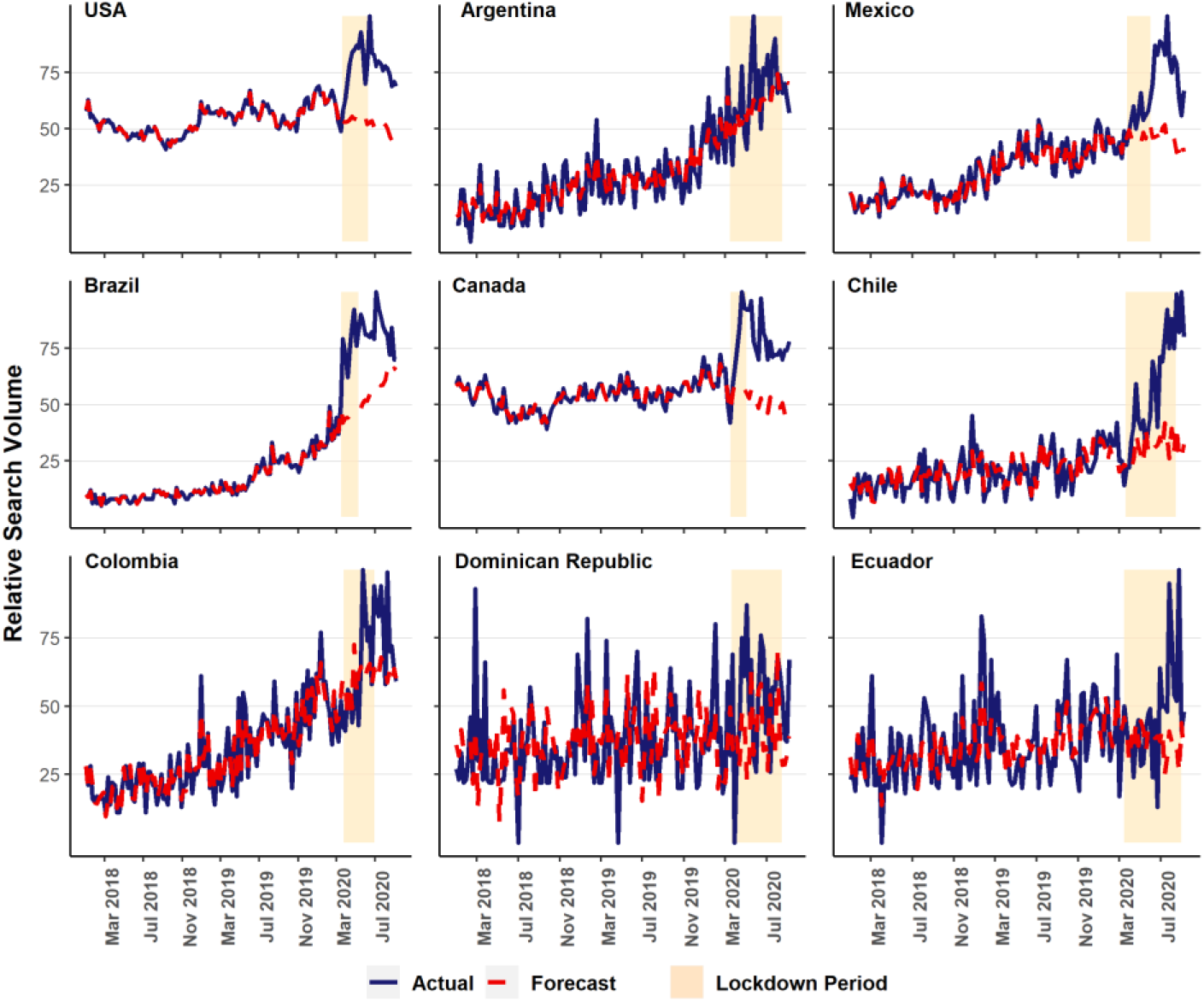
Acutal and ARIMA forecasted mean RSV values in American countries.

**Figure 3:**
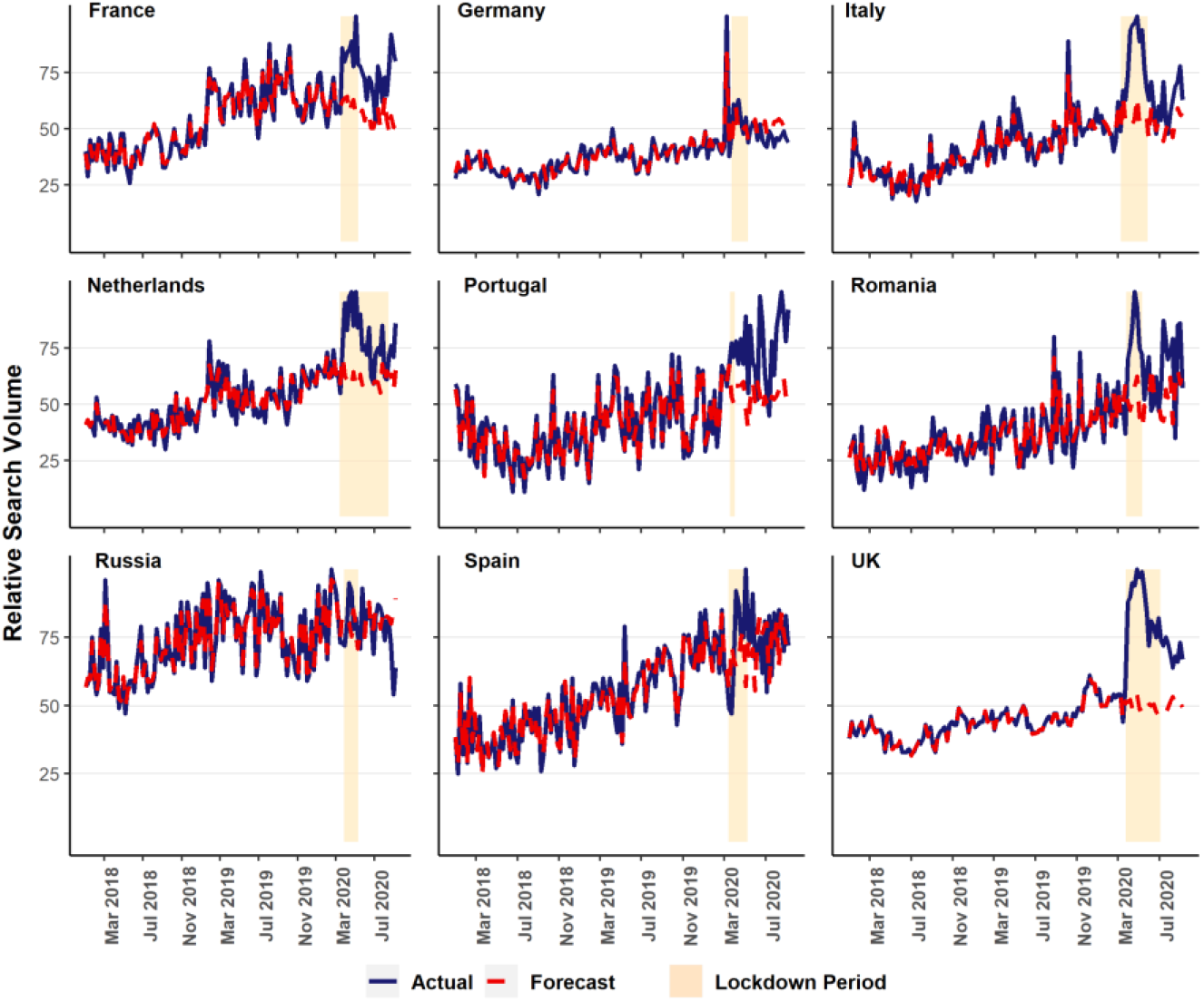
Acutal and ARIMA forecasted skincare mean RSV values in European countries.

## Discussion

In summary, the illustration of Google Trends data in the result section clearly shows that there is a significant change in the public interest in skincare treatment during the COVID-19 pandemic. Most of the selected countries showed a significant increment in Google Trends mostly in May, specifically, during the lockdown period. However, countries of Asia, the Dominican Republic, and Russia showed no significant change in public interests at all. These results suggest that people around the world have a concern about skincare treatment. Therefore, based on the increment in Google Trends, it can be assumed that people having skin related disease need skincare surveillance.

From the selected Americas countries, Chile showed the highest increment in RSVs for skincare in August (153.39%). This increment in Chile could be happened due to the lockdown (150 days) restriction which had seen to cause the sleeping problem, screen exposure, and disruption in behavioral appearance.^1,2^ Of all Americas countries, the USA was hit hard by most COVID-19 cases and deaths (WHO). This surge of cases caused a huge impact on the healthcare system in the USA such as in diabetics to care, substantial racial and ethnic disparities, and cognitive heart failure, etc.^25–27^ Like the other healthcare patients, the dermatology patients also affected by this consequence. More than 50% of the dermatology outpatients decreased in the USA during the lockdown period which might be a reason for the upward trend for skincare.^28^ After the USA, Brazil was the second country in the Americas which was mostly affected by COVID-19. The Google Trend for Brazil also followed the same way the USA went in the lockdown period.

The Google Trends for skincare in the UK showed the highest increment of all selected European countries. The trends started to increase when the lockdown was first imposed in the UK and reached the highest increment on April 26 (80.54%). A similar study showed that Google Trends for mental health deterioration also reached the highest increment in late April during the lockdown.^29^ This similarity in April suggests that the highest increment in the UK could be happened due to the lockdown measures. Besides the UK, this similar increment in trends was also found in Italy, France, and Belgium. In Italy, dermatological surgery patients in the restricted zone were reduced by approximately more than 45%.^30^ This decrement suggests the reason behind the continuous RSVs increment in Italy. Despite the surge of COVID-19 cases and deaths, Russia and Spain the only two countries showed little change in RSVs increment. However, this unexpected change in Russia may be related to Google’s lower search enginers martet share (< 60%) compared to other european countries.

From the analysis of Google Trends, it can be inferred that skincare outpatients have decreased during this COVID-19 pandemic. Therefore, people with skin problems need proper healthcare directions. For such a case, Telehealth might provide a probable solution to the skincare patients which is being widely used during this pandemic.^31,32^ The effectiveness of Telehealth has already been seen as patients surveillance in reducing the mental health burden, reducing exposure of obstetric patients, and surgical post-operative care, etc.^12,33–35^ Just like Telehealth, Teledermatogy could also provide patient surveillance for the skin disease patients.^36^ Besides, Teledermatogy would be a very useful service for those patients who are living in the rural side of the world.^37^ However, along with Telehealth, Teledermatogy has some limitations such as inadequate technical infrastructure, unequal reimbursement, and legal problems causing the delay of progress.^38,39^

In conclusion, this study provides an evaluation of public interests for skincare during this COVID-19 pandemic. The Google Trend analysis showed a significant increment in the public interest which could happen due to the lockdown restriction measures. The increment in public interests suggests the necessity of online contribution for skincare treatment. Since access to dermatologist care during lockdowns are scare and limited in most countries, the increasing public search interests in skin care also suggests the need for additional and urger medical care for untreated darmatological conditions. Finally, the analysis of Google Trends and the correlation between the COVID-19 pandemic and public interests for skincare can be help helpful for the dermatologists to analyze the present situation.

## Limitation

This study has some limitations. First, people who don’t have access to the internet excluded from this study which could bring significant change to Google Trends. However, since approximately 60% of the world population (4.57 billion people) are actively using the internet, it can be inferred that this population might be sufficient for skin disease patient’s surveillance.^40^ Second, this study was done only based on one search engine (Google Search Engine), the other popular search engines like Baidu in China, and Yandex in Russia can give a better estimation for public interests.^41,42^ Third, Google Trends ignores typographical error in the query terms which could also change the trend, especially, people with fewer skills could be excluded from this study for such a case. Finally, the COVID-19 cases and deaths are changing every day so at the time of doing this research, Google Trends can change significantly. In addition, countries around the world are now reopening some of their imposed restriction measures which might also bring some significant change in Google Trends.^43^ Further research is needed to explore the actual scenario causing the increase of public interest for skincare treatment.

## Data Availability

Data is obtained from Public Google Trends Website.

https://trends.google.com/trends/?geo=US

---

**Supplementary Table 1.**
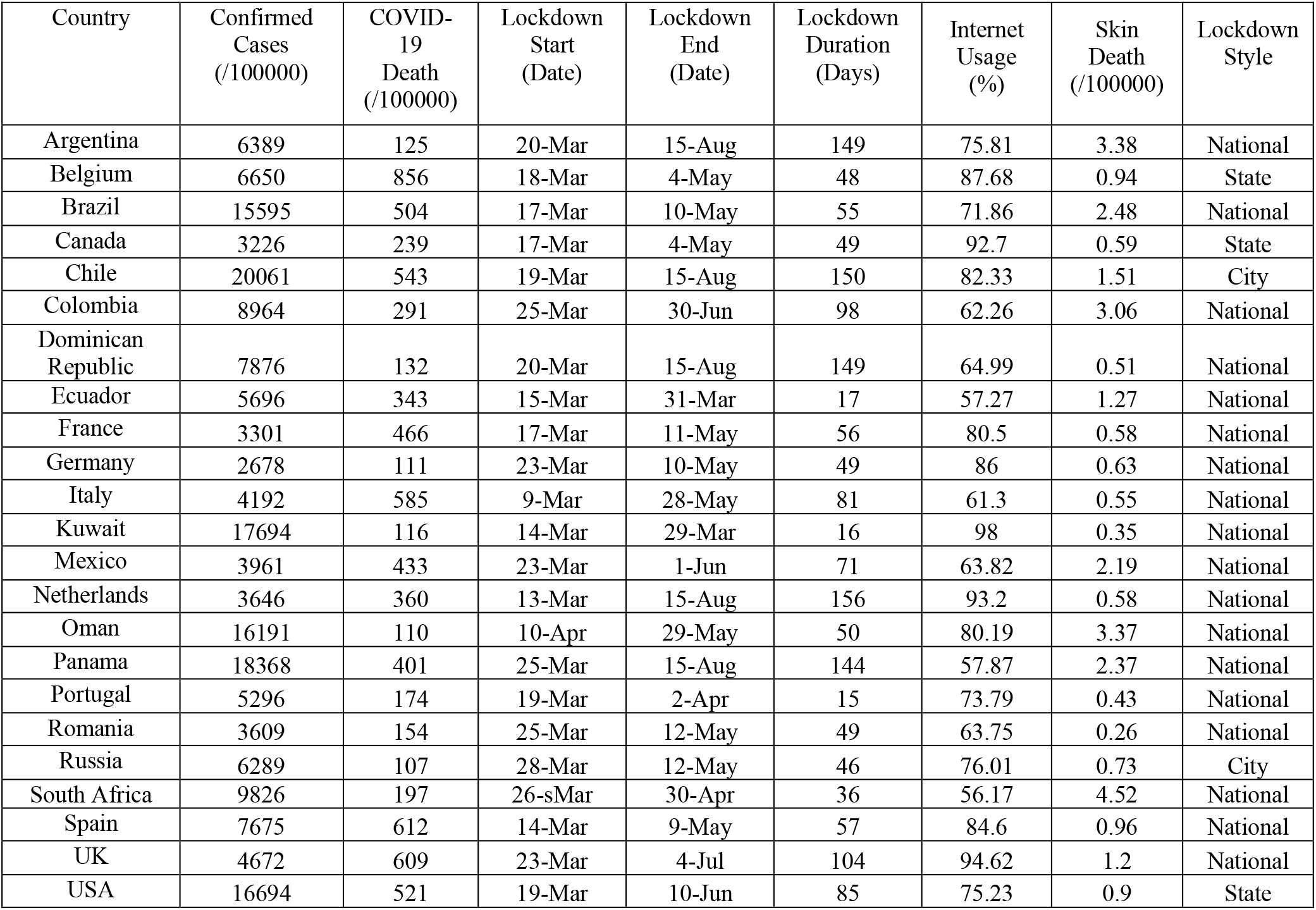
Characteristics of the individual countries during COVID-19 pandemic.

